# COVID-19 behavioural insights study: Preliminary findings from Finland, April-May, 2020

**DOI:** 10.1101/2020.10.11.20210724

**Authors:** Veronica Cristea, Timothee Dub, Oskari Luomala, Jonas Sivelä

## Abstract

The COVID-19 monitoring behavioural insights study was conducted from April-May 2020 in Finland. Respondents reported feeling confident protecting themselves against COVID-19 infection. Worries shifted from overloading the health system (mean value 5.5 [95% CI: 5.4-5.6]) to mental health concerns (mean value 5.3 [95% CI 5.2-5.4]). Maintaining physical distancing from families and friends decreased by 7% and 6%. Respondents mostly agreed that if a vaccine would become available, they would get it. The decrease in acceptance of recommended measures needs further analysis, but current results provide evidence to support the response.

**Key points:**

- Currently limited information available on the complex interaction between epidemiology, media attention, pandemic control measures, risk perception and compliance with public health measures.
- Despite the relatively high risk perception of a possible infection with COVID-19, we observed a steady decrease in adherence to public health measures.
- Throughout the study, information-seeking behaviour shifted.
- We observed a decrease in acceptance among the participants in regards to avoiding physical contact.

## Introduction

Previous studies have demonstrated that traditional media, social media and public health communication can influence knowledge, behaviour and perception of public health issues^1-2^ and influence preparedness and protective behaviour. ^3-4^Communication strategies should follow best practices for public health and risk communication and tailor messages accordingly, to address risk perceptions, attitudes toward risk avoidance, fears, worries and trust in authority^5^, and provide evidence-informed responses to misinformation, to support rational, adaptive and protective behaviours^6^. In April 2020, the Finnish Institute for Health and Welfare (THL) initiated regular monitoring of the Finnish population’s behaviour, knowledge, perception and trust regarding novel coronavirus disease (COVID-19). From April 7, 2020, we monitored these variables bi-weekly and shared findings with decision makers to help identify appropriate response measures, policies, interventions and communication strategies.

## Methods

We used the COVID-19 Snapshot Monitoring (COSMO Standard): Monitoring knowledge, risk perceptions, preventive behaviours, and public trust in the current coronavirus outbreak - WHO standard protocol^7-8^ to design a brief (approximately 10 minutes) online survey. Three iterative randomly-selected panels of approximately 1000 participants, representative of Finnish population in terms of age groups, gender and place of residence were invited to respond to items assessing risk perceptions, worries, fears, trust and information-seeking behaviour. We assessed risk perception as the perceived probability of contracting COVID-19, personal susceptibility and the expected severity of the disease using a 7 point Likert scale, from low to high, for which we calculated a mean value and 95% confidence intervals. Other question items addressed worries, trust in authorities, information-seeking and protective behaviours. Physical distancing behaviour was evaluated using closed questions with three possible responses: already done, planned or not planned at all. We used a 7 point Likert scale ranging from 1(disagree) to 7 (agree) to assess attitude towards a potential vaccine by asking participants how likely they would be to get vaccinated if a potential vaccine became available and recommended by Public Health authorities. We present preliminary findings of the first three rounds of data collection (Figure), 7-9 April (n= 1009), 24-28 April (n= 1032) and 8-11 May (n=1060).

**Figure.**
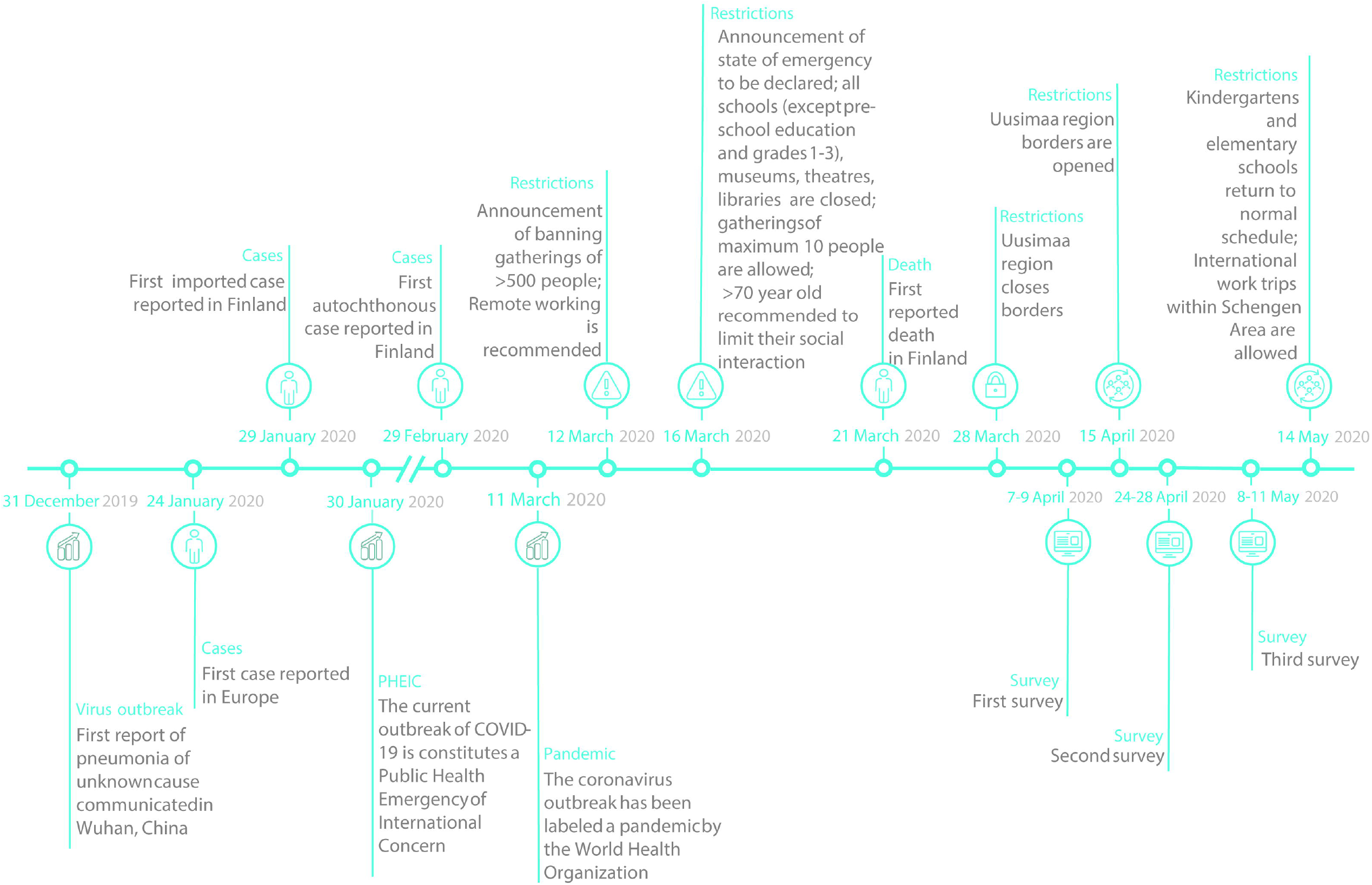
Timeline of coronavirus outbreak evolution and measures implemented in Finland, December 2019-May 2020.

## Results

### Risk perception, behaviour and trust

Respondents’ perception of their susceptibility to contracting COVID-19 was highest at the second time point, with a mean value of 3.8 (95% CI: 3.7-2.8). In the third round, perceived probability of contracting the disease and its perceived severity decreased to a mean of 3.4 (95% CI: 3.3-3.5) and 4.3 (95% CI: 4.2-4.4), respectively. Overall, respondents in the third round felt confident they could protect themselves against infection (mean value of 6 [95% CI: 5.92-6.05]).

At the first time point, 86% (864/1009) of participants had reduced their physical contacts with their families after the announcement of physical distancing recommendation on 12 March 2020. During the last round, 79% (835/1060) reported following this recommendation. Avoiding unnecessary social contact during the first time-point was reported by 86% (868/1009) compared to 80% (853/1060), at the third time-point.

Trust was evaluated using a 7 point scale from “very little trust” to “a great deal of trust”. Respondents in the third round scored a mean value of 4.7 (95% CI: 4.6-4.8) for the Ministry of Health, 4.9 (95% CI: 4.8-5) for THL and 4.7 (95% CI: 4.6-4.8) towards the government. Trust levels in different medias remained constant through the period, identifying television and online/ print news as the most trusted outlets, with mean values at the last time point of 5.0 (95% CI: 4.9-5.1) and 4.9 (95% CI: 4.8-5), compared to podcasts (mean value: 2.8 [95% CI: 2.7-2.9]) and social media (mean value: 2.7 [95% CI: 2.6-2.8]). A low level of trust in blogs was also evidenced with a mean value of 2.6 (95% CI: 2.4-2.7) at the last follow up.

### Worries

Throughout the whole study period, the coronavirus situation continued to worry and frighten Finnish respondents, but this appeared to be decreasing. In the first survey round, the leading worries were related to overloading of the health system (mean value 5.5 [95% CI: 5.4-5.6]), the situation of small companies (mean value 5.5 [95% CI: 5.4-5.6]) and economic recession (mean value 5.4 [95% CI: 5.3-5.5]), while during the third round, economic recession was the leading worry (mean value 5.3 [95% CI: 5.2-5.4]), followed by increased mental health concerns (mean value 5.3 [95% CI 5.2-5.4]) and the situation of small companies (mean value 5.3 [95% CI: 5.1-5.3]). Worries related to oneself (mean value 4 [95% CI: 3.9-4.1]) or of a loved person (mean value [95% CI: 4.6-4.8]) becoming infected were reported throughout the study period, but were not identified as the most important concerns.

### Acquisition of information and attitude towards the media

In the first round, participants reported searching COVID-19 related information often, with 40% (403/1009) declaring searching information multiple times a day, compared to 20% (214/1060) in the third round. Treatment progress development remained of interest during the whole study, with an increased interest in vaccine development during the third survey (mean value: 5.2 [95% CI: 5.1-5.3]) and a majority of respondents reporting willingness to be vaccinated against COVID-19 when opportunity arises (mean value 5.4 [95% CI: 5.3-5.5]).

### Gender differences

We assessed differences between male and female respondents, in terms of risk perception, worries, trust and information seeking patterns. Throughout the whole study period, women responded perceiving themselves to be more susceptible to contracting the infection, more confident in being able to protect themselves from infection, and more likely to avoid unnecessary physical contact (Supplementary material). A difference was observed in the leading worries, with males reporting lower levels of worry about overloading the health system, increasing mental health concerns, or infection of a person close to them during the whole time period. Measures of trust in different institutions over the two month period revealed that women were more trusting of the Ministry of Social Affairs and Health and the government.

Attitude towards a potential vaccine remained stable at all rounds. On a scale from 1(disagree) to 7(agree), mean value of how likely participants would get immunisation increased from 5.3 [95% CI: 5.2-5.4] to 5.4 [95% CI: 5.3-5.5] between the first and third survey round. Overall, most participants were in favor of receiving immunisation (69%, n=2141), 26% (n=809) mildly disagreed or had no opinion while 5% (n=151) strongly disagreed with benefiting from immunisation. There was no statistical difference regarding likelihood of receiving the vaccine between genders, however, age significantly influenced likelihood of receiving immunisation: the older respondents were, the most likely they were to get immunisation provided a vaccine would be available. This difference was evidenced at all time points. During the third wave, mean likelihood of getting immunisation ranged from 4.7[95% CI: 4.7-5.0] among 18-29 years old respondents to 6.1 [5.9-6.2] in respondents aged 65 years old or more.

## Discussion

This study provides evidence for decision makers and communication activities by assessing risk perceptions, worries, fears, trust in authority and information-seeking behavior during the COVID-19 epidemic, and describing changing attitudes over time, as the public health crisis develops. Currently, there is limited information available on the complex interaction between epidemiology, media attention, pandemic control measures, risk perception and compliance with public health measures.^9-10^Despite the relatively high risk perception of a possible infection with COVID-19, a steady decrease in adherence to public health measures was reported. Most respondents believed they are not highly susceptible to COVID-19, and if contracted, the disease would not be severe. Trust remained stable over the study period with worries expressed in different areas.

Surprisingly, own infection or infection of a loved one did not seem to be the greatest concern; rather, fears and worries were consistently related to economic recession, increase in mental health concerns and the situation of small businesses. Across the whole study, information-seeking behaviour shifted from searching information multiple times a day to several times a week. Measures such as avoiding physical contact with friends and family seem to be losing acceptance among the participants, indicating that such measures are becoming more difficult to sustain.

Overall attitude towards a potential vaccine was favorable, with older respondents more inclined to receive immunisation when it would become available.

As the COVID-19 epidemic evolves, it is important to understand the dynamics of risk perceptions, fears, misinformation and protective behaviours, identify which protective measures are employed, and what information is lacking. Based on continuing data collection and analysis, this tool makes it possible for authorities to react quickly to emerging misinformation or sudden increases in risk perceptions that could foment panic.

## Supporting information

Risk perceptions, trust, worries and information acquisition responses by gender, Finland, April- May 2020

## Data Availability

Data can be made available upon request to the authors

## Funding

This study was funded by the Finnish Institute for Health and Welfare.

## Conflicts of interest

None to declare.

## Acknowledgments

We would like to acknowledge numerous persons that contributed to developing of the COVID-19 Snapshot Monitoring (COSMO Standard): Monitoring knowledge, risk perceptions, preventive behaviours, and public trust in the current coronavirus outbreak protocol and guidance: Cornelia Betsch (PI), Lars Korn, Lisa Felgendreff, Sarah Eitze, Philipp Schmid, Philipp Sprengholz from Universität Erfurt, Katrine Bach Habersaat and Martha Scherzer from WHO Regional Office for Europe; Taloustutkimus Oy team for input on questionnaire design, implementation and data collection; Idil Hussein for her input during the questionnaire design, Lotta Siira for input regarding the timeline of the events and Mika Salminen, Jussi Sane, Taneli Puumalainen, Katja Sibenberg from the Finnish Institute for Health and Welfare for their support and collaboration. Furthermore, we thank Outi Lyytikäinen and Lisa Hansen for their constructive comments on the manuscript.

## Notes

### Competing Interest Statement

The authors have declared no competing interest.

### Funding Statement

This work was funded by the Finnish institute for Health and Welfare

### Author Declarations

The steering committee of the Infectious Disease Control and Vaccinations of the Department of Health Security at the Finnish institute for Health and Welfare exempted this research from further ethical review.

