## Supplementary material for "COVID-19 behavioural insights study: Preliminary findings from Finland, April-May, 2020": Risk perceptions, trust, worries and information acquisition responses by gender, Finland, April- May 2020

### TABLE

Risk perceptions, trust, worries and information acquisition responses by gender, Finland, April- May 2020.

|  |  | All participants^a^ | | | Female | | | Male | | |  | | |
| --- | --- | --- | --- | --- | --- | --- | --- | --- | --- | --- | --- | --- | --- |
|  | Survey | n | Mean | SD | n | Mean | SD | n | Mean | SD | *df* | T | Significance |
| How susceptible do you consider yourself to an infection | 7-9 April | 986 | 3.73 | 1.41 | 502 | 3.95 | 1.44 | 482 | 3.49 | 1.35 | 3.99 | 4.59 | 0.01^b^ |
|  | 24-28 April | 1014 | 3.75 | 1.47 | 509 | 3.91 | 1.52 | 503 | 3.58 | 1.40 | 3.98 | 3.17 | 0.03 ^b^ |
|  | 8-11 May | 1039 | 3.61 | 1.43 | 535 | 3.79 | 1.51 | 502 | 3.43 | 1.32 | 3.96 | 3.58 | 0.02 ^b^ |
| How probable is it for you to contract an infection | 7-9 April | 986 | 3.52 | 1.48 | 502 | 3.67 | 1.47 | 482 | 3.36 | 1.47 | 3.99 | 2.93 | 0.04 ^b^ |
|  | 24-28 April | 1014 | 3.47 | 1.48 | 509 | 3.53 | 1.46 | 503 | 3.42 | 1.50 | 3.99 | 1.04 | 0.35 |
|  | 8-11 May | 1039 | 3.38 | 1.45 | 535 | 3.48 | 1.40 | 502 | 3.27 | 1.51 | 3.99 | 2.04 | 0.11 |
| Severity of the infection | 7-9 April | 986 | 4.49 | 1.57 | 502 | 4.62 | 1.51 | 482 | 4.37 | 1.63 | 3.96 | 2.18 | 0.09 |
|  | 24-28 April | 1014 | 4.39 | 1.67 | 509 | 4.56 | 1.60 | 503 | 4.23 | 1.72 | 3.97 | 2.76 | 0.05 ^b^ |
|  | 8-11 May | 1039 | 4.34 | 1.58 | 502 | 4.22 | 1.61 | 502 | 4.45 | 1.55 | 3.98 | 2.09 | 0.10 |
| Feeling prepared to avoid an infection | 7-9 April | 1009 | 5.95 | 1.08 | 510 | 6.17 | 0.97 | 497 | 5.73 | 1.14 | 3.80 | 6.11 | 0.004 ^b^ |
|  | 24-28 April | 1032 | 5.98 | 1.05 | 511 | 6.23 | 0.91 | 519 | 5.74 | 1.12 | 3.88 | 6.46 | 0.003 ^b^ |
|  | 8-11 May | 1060 | 6.00 | 1.08 | 545 | 6.22 | 0.92 | 513 | 5.78 | 1.18 | 3.77 | 5.86 | 0.005 ^b^ |
| Feeling able to avoid an infection | 7-9 April | 1009 | 5.01 | 1.41 | 510 | 5.06 | 1.48 | 497 | 4.97 | 1.33 | 3.96 | 0.94 | 0.39 |
|  | 24-28 April | 1032 | 5.00 | 1.41 | 511 | 5.01 | 1.47 | 519 | 4.98 | 1.35 | 3.97 | 0.25 | 0.81 |
|  | 8-11 May | 1060 | 5.02 | 1.41 | 545 | 5.05 | 1.45 | 513 | 4.99 | 1.36 | 3.99 | 0.60 | 0.58 |
| Avoiding unnecessary physical contact | 7-9 April | 1009 | 1.22 | 0.58 | 510 | 1.13 | 0.45 | 497 | 1.31 | 0.67 | 3.48 | -4.35 | 0.01 ^b^ |
|  | 24-28 April | 1032 | 1.28 | 0.66 | 511 | 1.19 | 0.57 | 519 | 1.36 | 0.72 | 3.82 | -3.71 | 0.02 ^b^ |
|  | 8-11 May | 1060 | 1.33 | 0.71 | 545 | 1.19 | 0.56 | 513 | 1.49 | 0.81 | 3.48 | -6.24 | 0.005 ^b^ |
| Overloading the health system | 7-9 April | 1009 | 5.48 | 1.36 | 510 | 5.69 | 1.28 | 497 | 5.28 | 1.42 | 3.94 | 4.29 | 0.01 ^b^ |
|  | 24-28 April | 1032 | 5.00 | 1.38 | 511 | 5.29 | 1.27 | 519 | 4.71 | 1.43 | 3.95 | 6.08 | 0.004 ^b^ |
|  | 8-11 May | 1060 | 4.93 | 1.41 | 545 | 5.16 | 1.35 | 513 | 4.69 | 1.43 | 3.96 | 4.83 | 0.009 ^b^ |
| Worries of small companies situation | 7-9 April | 1009 | 5.50 | 1.42 | 510 | 5.62 | 1.36 | 497 | 5.38 | 1.47 | 3.96 | 2.35 | 0.08 |
|  | 24-28 April | 1032 | 5.31 | 1.37 | 511 | 5.43 | 1.33 | 519 | 5.18 | 1.41 | 3.99 | 2.57 | 0.06 |
|  | 8-11 May | 1060 | 5.26 | 1.42 | 545 | 5.45 | 1.33 | 513 | 5.05 | 1.49 | 3.92 | 4.00 | 0.02 ^b^ |
| Worries of recession | 7-9 April | 1009 | 5.43 | 1.51 | 510 | 5.51 | 1.52 | 497 | 5.36 | 1.50 | 4.00 | 1.38 | 0.23 |
|  | 24-28 April | 1032 | 5.32 | 1.49 | 511 | 5.45 | 1.42 | 519 | 5.19 | 1.56 | 3.97 | 2.48 | 0.07 |
|  | 8-11 May | 1060 | 5.33 | 1.51 | 545 | 5.48 | 1.40 | 513 | 5.17 | 1.61 | 3.88 | 2.99 | 0.04 ^b^ |
| Worries of increase in mental health concerns | 7-9 April | 1009 | 5.20 | 1.38 | 510 | 5.41 | 1.31 | 497 | 4.98 | 1.42 | 3.96 | 4.40 | 0.01 ^b^ |
|  | 24-28 April | 1032 | 5.14 | 1.36 | 511 | 5.41 | 1.28 | 519 | 4.86 | 1.38 | 3.98 | 5.88 | 0.004 ^b^ |
|  | 8-11 May | 1060 | 5.29 | 1.35 | 545 | 5.59 | 1.20 | 513 | 4.98 | 1.43 | 3.84 | 6.61 | 0.003^b^ |
| Worries regarding ones’ own possibility of infection | 7-9 April | 1009 | 4.38 | 1.52 | 510 | 4.60 | 1.57 | 497 | 4.16 | 1.42 | 3.97 | 4.07 | 0.02 ^b^ |
|  | 24-28 April | 1032 | 4.14 | 1.51 | 511 | 4.36 | 1.51 | 519 | 3.92 | 1.48 | 3.99 | 4.19 | 0.01 ^b^ |
|  | 8-11 May | 1060 | 4.09 | 1.56 | 545 | 4.25 | 1.58 | 513 | 3.93 | 1.51 | 3.99 | 2.96 | 0.04 |
| Worries regarding someone close getting infected | 7-9 April | 1009 | 4.97 | 1.49 | 510 | 5.22 | 1.51 | 497 | 4.72 | 1.42 | 3.99 | 4.77 | 0.009 ^b^ |
|  | 24-28 April | 1032 | 4.78 | 1.49 | 511 | 5.05 | 1.48 | 519 | 4.52 | 1.46 | 3.99 | 5.15 | 0.007 ^b^ |
|  | 8-11 May | 1060 | 4.74 | 1.57 | 545 | 4.94 | 1.53 | 513 | 4.51 | 1.59 | 3.98 | 3.96 | 0.01 ^b^ |
| Trust the Ministry of Health | 7-9 April | 969 | 4.90 | 1.50 | 493 | 5.15 | 1.31 | 474 | 4.64 | 1.63 | 3.80 | 4.71 | 0.01 ^b^ |
|  | 24-28 April | 997 | 4.63 | 1.55 | 489 | 4.81 | 1.49 | 506 | 4.46 | 1.59 | 3.99 | 3.14 | 0.03 ^b^ |
|  | 8-11 May | 1040 | 4.71 | 1.53 | 531 | 4.94 | 1.39 | 507 | 4.46 | 1.64 | 3.87 | 4.46 | 0.01 ^b^ |
| Trust the Finnish Institute for Health and Welfare | 7-9 April | 988 | 4.99 | 1.65 | 504 | 5.31 | 1.43 | 482 | 4.67 | 1.79 | 3.78 | 5.43 | 0.007 ^b^ |
|  | 24-28 April | 1012 | 4.82 | 1.58 | 500 | 4.93 | 1.54 | 510 | 4.71 | 1.61 | 3.99 | 1.99 | 0.11 |
|  | 8-11 May | 1043 | 4.93 | 1.60 | 534 | 5.17 | 1.45 | 507 | 4.67 | 1.71 | 3.86 | 4.47 | 0.01 ^b^ |
| Trust the government | 7-9 April | 982 | 4.94 | 1.54 | 500 | 5.24 | 1.33 | 480 | 4.64 | 1.68 | 3.76 | 5.47 | 0.006 ^b^ |
|  | 24-28 April | 1015 | 4.74 | 1.61 | 500 | 4.90 | 1.54 | 513 | 4.59 | 1.66 | 3.98 | 2.76 | 0.05 ^b^ |
|  | 8-11 May | 1044 | 4.74 | 1.63 | 534 | 5.00 | 1.46 | 508 | 4.47 | 1.75 | 3.84 | 4.67 | 0.01 ^b^ |
| Frequency of information search | 7-9 April | 1009 | 2.12 | 1.30 | 510 | 2.03 | 1.29 | 497 | 2.22 | 1.31 | 3.99 | -2.02 | 0.11 |
|  | 24-28 April | 1032 | 2.41 | 1.35 | 511 | 2.24 | 1.24 | 519 | 2.56 | 1.43 | 3.93 | -3.38 | 0.02 ^b^ |
|  | 8-11 May | 1060 | 2.77 | 1.43 | 545 | 2.64 | 1.37 | 513 | 2.89 | 1.47 | 3.96 | -2.52 | 0.06 |
| Information regarding vaccine development progress is needed | 7-9 April | 992 | 5.02 | 1.49 | 504 | 5.13 | 1.48 | 486 | 4.91 | 1.50 | 3.99 | 2.08 | 0.10 |
|  | 24-28 April | 1014 | 4.99 | 1.53 | 500 | 5.12 | 1.53 | 512 | 4.88 | 1.52 | 3.99 | 2.19 | 0.09 |
|  | 8-11 May | 1039 | 5.20 | 1.50 | 537 | 5.33 | 1.43 | 500 | 5.05 | 1.55 | 3.94 | 2.65 | 0.05 |
| If a vaccine is available and recommended to me I will take it | 7-9 April | 1009 | 5.30 | 1.70 | 510 | 5.31 | 1.73 | 497 | 5.30 | 1.67 | 3.99 | 0.15 | 0.88 |
|  | 24-28 April | 1032 | 5.26 | 1.76 | 511 | 5.24 | 1.81 | 519 | 5.28 | 1.72 | 3.98 | -0.31 | 0.77 |
|  | 8-11 May | 1060 | 5.39 | 1.74 | 545 | 5.39 | 1.76 | 513 | 5.39 | 1.74 | 3.99 | -0.05 | 0.95 |

^a^ Data displayed includes female, male and other

^b^ Significant (p-value ≤0.05) using Welch’s two-sample t test
